# Resting-state brain function and its modulation by intranasal oxytocin in antisocial personality disorder with and without psychopathy

**DOI:** 10.1101/2025.04.08.25325470

**Authors:** Julia Griem, Daniel Martins, John Tully, Declan Murphy, Yannis Paloyelis, Nigel Blackwood

**Affiliations:** Department of Clinical, Educational and Health Psychology, Division of Psychology and Language Sciences, University College London, UK; Department of Forensic and Neurodevelopmental Sciences, Institute of Psychiatry, Psychology and Neuroscience, King’s College London, UK; Anna Freud, London, UK; Department of Neuroimaging, Institute of Psychiatry, Psychology and Neuroscience, King’s College London, UK; Academic Unit of Mental Health and Clinical Neurosciences, School of Medicine, Institute of Mental Health, University of Nottingham, UK

## Abstract

Behavioural, structural and functional neuroimaging differences have been demonstrated between individuals with antisocial personality disorder with (ASPD+P) or without psychopathy (ASPD-P). However, the underlying mechanisms for such differences are poorly understood, hampering progress in the development of drug treatments for this population. Intranasal oxytocin (OT) has garnered significant attention due to its prosocial effects in healthy individuals. We sought to establish the impact of OT on resting-state brain function in individuals with ASPD, and to explore whether modulation differs between individuals with and without psychopathy. We used arterial spin labelling (ASL) to measure regional cerebral blood flow (rCBF) to investigate brain function at rest and modulation of key disease-targets by a single acute dose of OT (40 IU). We used a double-blind, placebo-controlled, crossover design in males with a history of violent offending with ASPD+P (N = 17) or ASPD-P (N = 14) and a group of healthy male non-offenders (N = 22). Both ASPD subtypes showed reduced rCBF in frontotemporal regions compared to non-offenders. However, those with ASPD+P demonstrated significantly greater rCBF increases in posterior default mode network regions compared to those with ASPD-P. OT administration selectively reduced rCBF in the left basal ganglia of the ASPD-P group, an effect not observed in the ASPD+P or non-offender groups. Our results provide further evidence of functional brain differences between ASPD+P and ASPD-P groups, and a differential modulating effect of oxytocin. The neurobiological distinctions between ASPD+P and ASPD-P groups are important considerations for future therapeutic developments.

## Introduction

Antisocial personality disorder (ASPD) is characterized by impulsivity, irresponsibility and aggression [1]. Approximately one third of individuals with ASPD additionally meet categorical criteria for psychopathy (ASPD+P) [2], characterized by callous unemotional behaviours and a lack of remorse [3]. While individuals with and without (ASPD-P) psychopathy share features such as life-course persistent offending, heightened reactive aggression, and cognitive impairments [4–6], there are also differences between the groups: those with ASPD+P demonstrate an earlier onset and greater density of offending behaviours, increased use of proactive aggression, and a poorer response to interventions developed for offending populations [7–10]. Such behavioural abnormalities are associated with shared and distinct structural and functional brain differences in networks associated with reward and empathic processing such as the dorsomedial prefrontal cortex, striatum, insula, cingulate cortex and amygdala [11–16].

While prior functional studies in these populations have focused on task-related brain function in ASPD, resting-state analysis offers unique insights into baseline neural activity independent of specific behavioural demands [17–20]. Resting-state function is linked to, and may even predict, general cognitive propensities that underpin behaviour such as decision-making [21– 23]. Studies investigating resting-state brain activity in large samples of imprisoned offenders with varying degrees of psychopathy have suggested abnormalities in the form of aberrant functional connectivity or topology in frontotemporal and limbic regions including the anterior cingulate, insula, and amygdala, as well as in large-scale brain networks including the default mode network (DMN) [24–26]. However, no study to date has assessed similarities or differences in resting-state brain activity between ASPD+P and ASPD-P populations.

In addition, resting-state imaging studies in these populations have typically relied on measurements of the blood-oxygen-level-dependent (BOLD) contrast, a proxy measure of neural activity based on the complex interaction between blood flow, volume and oxygen metabolism [27,28]. Regional cerebral blood flow (rCBF) approaches provide a more direct physiological proxy of neural activity by indexing perfusion from cerebral capillaries into brain tissue. A small number of studies have measured rCBF using single photon emission computed tomography (SPECT) or positron emission tomography (PET) in antisocial populations, typically demonstrating decreased perfusion in anterior and medial (cingulate) frontal regions, lateral temporal regions, the amygdala and insula [29,30], and increased perfusion in posterior DMN regions [31]. However, these studies were constrained by the recruitment of heterogenous clinical and non-clinical samples, including individuals with aggression, antisocial behaviours or substance misuse disorders, but without detailed phenotypic characterization of ASPD+/-P. Furthermore, these studies used large regions of interest and non-automated analytic approaches to perfusion data. These limitations can be addressed by a more recently developed imaging approach, arterial spin labelling (ASL), which offers an endogenous, non-invasive measure of rCBF, with improved spatial resolution and excellent test-retest reliability [32,33]. No study to date has used ASL to assess resting-state rCBF in individuals with carefully phenotyped ASPD+/-P.

Measurement of resting-state brain activity in clinical populations also offers a context-free measure of the brain’s responsivity to pharmacological modulation. One pharmacological agent of particular interest to the ASPD+/-P populations is oxytocin, a neuropeptide central to the regulation of complex social behaviours such as empathy [34,35]. Two recent studies utilising BOLD imaging techniques revealed that a single dose of intranasal oxytocin (OT) normalised aberrant neural responsivity to emotional stimuli in the anterior cingulate and anterior insula in the ASPD+P group [36] and the amygdala in a broadly defined ASPD group [37].

ASL imaging is more sensitive to the effects of pharmacological challenges than BOLD imaging because rCBF is less susceptible to non-specific drug effects than the BOLD signal [28,38–40]. Its ability to capture the time-, method-, and dose-dependent changes in rCBF in response to oxytocin has been demonstrated in healthy individuals [41,42]. Intranasal OT modulates rCBF in regions including the amygdala, anterior insula, and striatum [42,43], where dysregulated neural activity has been demonstrated in ASPD+/-P populations [13,44]. However, whether OT also modulates resting-state brain function (rCBF) in ASPD, and importantly whether this modulation is moderated by the presence of psychopathy remains unknown.

To address these outstanding questions, we employed ASL imaging to examine the impact of a single acute intranasal administration of OT on resting-state rCBF in males with a history of violent offending with ASPD+P and ASPD-P and healthy male non-offending controls using a double-blind, placebo-controlled, randomised crossover design. We conducted whole-brain and region-of-interest (ROIs: amygdala/anterior insula) analyses. For the whole-brain analyses, we hypothesized that: (1) both ASPD groups would show reduced anterior frontal, anterior cingulate and lateral temporal lobe rCBF, together with increased rCBF in posterior DMN regions such as posterior cingulate and precuneus in comparison to the non-offender group and (2) such differences would be more marked in the ASPD+P group in comparison to the ASPD-P group. For the ROI analysis, we hypothesized that: (3) both ASPD groups would show reduced amygdala and anterior insula rCBF in comparison to the non-offender group; and (4) such differences would be more marked in the ASPD+P group in comparison to the ASPD-P group Finally, (5) OT would restore rCBF in frontotemporal regions in ASPD+P but not ASPD-P. We also explored the potential functional impact of any identified differences by correlating the imaging findings with phenotypic characteristics of the violent offenders such as recidivism rates.

## Materials and methods

### Participants

Between September 2017 and March 2020, we enrolled 53 male participants (22 healthy non-offenders and 31 offenders with ASPD with (N=17) or without (N=14) psychopathy) aged 18-60, with normal range IQ according to the Wechsler Abbreviated Scale of Intelligence (WASI-II) [45]. We recruited offenders with convictions for violent crimes (murder, rape, attempted murder, grievous and actual bodily harm) who met DSM-5 criteria for ASPD via the National Probation Service of England and Wales and local forensic personality disorder services. We recruited healthy non-offenders from the general population through public and online advertising. All participants completed diagnostic interviews (Structured Clinical Interview for the DSM-5-Clinical and Personality Disorders (SCID-5-CV/SCID-5-PD) [1,46] and Psychopathy Checklist-Revised (PCL-R) [3]) and authorized access to their criminal records. In line with previous research in UK samples [11,13,36,44,47], we used a score of 25 as the threshold for psychopathy in this English population. We excluded participants if they had a history of major mental illness (bipolar 1, bipolar 2, major depression or psychotic disorders), self-reported neurological disorders, head injury resulting in loss of consciousness for 1h or longer, severe visual or hearing impairments, or contraindications to MRI.

The study was approved by London City and East Research Ethics Committee (15/LO/1083), as well as the National Offender Management Services Research Committee (2016-382). After receiving a complete description of the study, all participants completed signed consent. All assessments were conducted by an experienced research psychologist (JG) and forensic psychiatrist (JT). This trial was registered at ClinicalTrials.gov (ID NCT05383300). Participants completed the self-report Reactive-Proactive Aggression Questionnaire [48]. On the day of each MRI scan, participants provided a urine sample to assess for substance use.

### Study design and procedure

We used a double-blind, placebo-controlled, randomised crossover design. We acquired anatomical and ASL scans as part of a larger imaging protocol (see supplementary materials for the full schedule, noting the gap between task-based scans and ASL imaging to minimise carry-over effects). For the two scanning sessions, we randomly and blindly allocated the participant to receive 40 international units (IU) of OT (Syntocinon, Novartis, Switzerland) or placebo (PL; same excipient without the oxytocin). Scans were scheduled at least 3 days apart to ensure complete drug washout. We counterbalanced the order of administration for OT/PL across participants, with half receiving OT first and then PL, and the other half receiving the reverse order. According to the recommendation for standardised administration [49] and under supervision of the researcher, participants received training and then self-administered the nasal spray by snorting one puff every 30 seconds through alternating nostrils, for 5 minutes (10 puffs with 4 IU each). We selected the oxytocin dose in line with safety standards and previous studies measuring effects of OT on rCBF [41–43,50]. This dose was not associated with any side effects. The ASL scans (6:23 minutes) were acquired on average at 84 (±9) minutes and 85 (±12) minutes after administration of PL and OT, respectively. This time delay is referred to as the variable ‘minutes since dose’. Scans were acquired at the same time of the day within and between participants (see supplementary materials for details).

### Image acquisition

We used a General Electric MR750 3Tesla MRI scanner and 32-channel C-RMNova head coil for this study. During each session, we acquired a 3-dimensional pseudo-continuous ASL scan (60 slice partitions with thickness and gap = 3 mm, TE = 1109 ms, TR = 5180 ms, flip angle = 111°, FOV = 240 x 240 mm^2^, in-plane resolution = 3.6 mm; radiofrequency inversion pulse = 1825 ms, delay = 2025 ms, control-label image pairs = 5). The acquisition of the final rCBF map for each participant at each session was in line with recommendations [51].

We also acquired a 3D high-resolution T1-weighted whole-brain anatomical image during each scanning session (MPRAGE, 196 slices with thickness and gap = 1.2 mm, TE = 3.02 ms, TR = 7.31 ms, TI = 400 ms, FA = 11°, FOV = 270 x 270 mm^2^, matrix = 256 x 256, voxel resolution = 1.05 x 1.05 x 1.2 mm^3^).

### Image pre-processing

We confirmed good image quality and presence of typical perfusion values ranging between 20 and 110 ml/100 g/minute across the whole brain, indicating accurate computation of CBF maps [51], using FSLeyes [52]. We pre-processed the scans in the Automatic Software for ASL Processing (ASAP) toolbox, version 4.0 in Matlab 2018b [53]. Steps included (1) co-registration of proton density and T1 images; (2) subject-specific normalization of CBF maps; (3) skull-stripping, segmentation and removal of extra-cerebral signal from the normalized CBF maps; (4) partial volume correction and normalization of the CBF map to MNI152 space; and (5) 8 mm Gaussian spatial smoothing of the CBF map. An explicit grey matter tissue probability mask (20%), derived by thresholding the FSL grey matter template at 0.20, was applied. We calculated global median CBF for each participant and each session and included this in statistical analysis as a covariate of no-interest to improve signal-to-noise ratio and the sensitivity of within-subject changes in local areas. We used median rCBF values throughout due to a skew towards the lower end of the typical range.

### Statistical analysis

We compared demographic and clinical characteristics, as well as global median CBF using ANOVA, ANCOVA, and Chi-squared tests, or non-parametric equivalents when normality assumptions were not met, in SPSSv29 [54]. We interpreted significant main effects via Bonferroni-corrected pairwise comparisons.

We conducted whole-brain analyses in SPM12 (www.fil.ion.ucl.ac.uk/spm) using a partitioned errors approach to accommodate repeated-measures design assumptions [55]. We examined the main effect of group using a one-way ANOVA on individual-averaged OT and PL CBF maps. We assessed the main effect of treatment via a one-sample t-test using difference images, subtracting an individual’s PL CBF map from their OT CBF map. We tested the group by treatment interaction effect with a one-way ANOVA on the difference images. We covaried for global CBF, age, and minutes since dose in all models. We applied F-contrasts to assess main and interaction effects, with cluster-based inference using a cluster-forming threshold of p = 0.005 and familywise error correction at α = 0.05; in accordance with prior ASL studies investigating OT effects [42,43,56]. For significant clusters, we extracted and compared raw median CBF values using post-hoc pairwise comparisons or simple main effects tests with the Sidak correction for multiple comparisons in SPSSv29.

For the ROI analysis, we extracted median rCBF from the bilateral amygdala and anterior insula using Harvard-Oxford atlas masks in MNI space. We assessed main effects of group, treatment, and their interaction with bootstrapped linear mixed models in JASP [57], covarying for global CBF, age, and minutes since dose. We applied FDR correction for multiple comparisons across the ROIs. To further evaluate the robustness of our findings, we conducted Bayesian linear mixed models post hoc (see Supplementary Methods).

In ASPD participants, we completed exploratory bootstrapped partial correlation analyses (SPSSv29) to examine associations between median rCBF in regions showing significant group, treatment, or interaction effects and phenotypic characteristics (number of violent convictions, the presence of a reconviction within 3 years of participation, PCL-R factor scores, reactive and proactive aggression). These correlations accounted for group, global CBF, age, and minutes since dose, with multiple comparisons controlled using FDR correction.

## Results

### Sample characteristics

Table 1 shows the demographic and clinical characteristics of the three participant groups. As expected, the three groups differed significantly in years of education (offenders had fewer years of education than non-offenders), PCL-R scores, and the aggression subscales (offenders scored higher than non-offenders, and the ASPD+P group scored higher than the ASPD-P group). They did not significantly differ on age or IQ. Also as expected, the ASPD+P group had a higher frequency of comorbid cluster A personality disorders than the ASPD-P group. This is in keeping with the normal range of variation in clinical profiles of ASPD+/-P and we did not adjust our analyses based on these findings. Finally, the two ASPD groups did not significantly differ in comorbid lifetime substance use disorder or positive urine drug screening tests on the day of testing. To avoid over-correcting for phenotypic variance inherent to ASPD [58], substance use was not included as a covariate (see supplementary materials for results that included this as a covariate). Global CBF did not differ between groups or treatment conditions (see supplementary table 1).

**Table 1.** Demographic and clinical characteristics of participants. Note: Data are mean (standard deviation) unless otherwise stated. Non-offender participants did not have any diagnosis of personality disorder or mental illness and no history of conviction, therefore only the two ASPD groups were compared on these variables. Some participants did not complete the RPQ (final ASPD+P N = 13, ASPD-P N = 13, NO N = 20). RPQ = reactive proactive aggression questionnaire, PD = personality disorder, SUD = substance use disorder. F-statistic = ANOVA with Tukey post-hoc, H-statistic = Kruskal Wallis with Mann Whitney U post hoc, ^†^ Fisher’s Exact test with Fisher’s Exact post-hoc, χ^2^ = chi-squared test of independence with chi-squared test of independence as post hoc, U-statistic = Mann Whitney U. **◊** pairwise comparison did not survive a Bonferroni correction for multiple comparisons (α= .05/3 = .02).

| Demographic | ASPD+P<br>(N = 17) | ASPD-P<br>(N = 14) | NO<br>(N = 22) | Group comparison | Post hoc tests (p-values) |  |  |
| --- | --- | --- | --- | --- | --- | --- | --- |
|  |  |  |  |  | NO vs<br>ASPD+P | NO vs<br>ASPD-P | ASPD+P<br>vs<br>ASPD-P |
| Age (years) | 40.56 (9.95) | 45.21 (10.05) | 37.73 (9.56) | $H_{(2)} = 4.44, p = 0.11$ | . | . | . |
| IQ | 91.56 (12.45) | 99.14 (15.24) | 98.95 (10.77) | $H_{(2)} = 3.44, p = 0.18$ | . | . | . |
| Duration of education (years) | 9.88 (1.82) | 10.64 (2.21) | 13.77 (3.27) | $H_{(2)} = 19.40, p < 0.001$ | $p < 0.001$ | $p = 0.002$ | $p = 0.28$ |
| Age at first violent conviction | 20.38 (5.45) | 21.43 (5.03) | . | $t = 0.55, p = 0.59$ | . | . | . |
| Violent convictions | 4.31 (2.87) | 3.86 (2.93) | . | $U = 112.00, p = 0.78$ | . | . | . |
| Reconviction within 3 years, N (%) | 8 (47%) | 6 (43%) | . | $0.06, p = 1.00$ † | . | . | . |
| RPQ reactive aggression | 15.77 (4.69) | 12.46 (5.77) | 5.90 (3.54) | $F_{(2)} = 19.90, p < 0.001$ | $p < 0.001$ | $p < 0.001$ | $p = 0.22$ |
| RPQ proactive aggression | 13.85 (6.39) | 7.38 (6.41) | 0.75 (1.16) | $H_{(2)} = 29.30, p < 0.001$ | $p < 0.001$ | $p < 0.001$ | $p = 0.03^{\diamond}$ |
| RPQ total aggression | 29.62 (9.91) | 19.85 (11.46) | 6.65 (4.03) | $H_{(2)} = 27.56, p < 0.001$ | $p < 0.001$ | $p < 0.001$ | $p = 0.04^{\diamond}$ |
| PCL-R Factor 1 | 9.24 (3.05) | 4.81 (3.03) | 1.09 (1.63) | $H_{(2)} = 35.41, p < 0.001$ | $p < 0.001$ | $p < 0.001$ | $p < 0.001$ |
| PCL-R Facet 1 (interpersonal) | 4.24 (1.82) | 1.81 (1.63) | 0.64 (0.95) | $H_{(2)} = 27.80, p < 0.001$ | $p < 0.001$ | $p = 0.02$ | $p < 0.001$ |
| PCL-R Facet 2 (affective) | 5.00 (1.80) | 3.00 (1.80) | 0.46 (0.80) | $H_{(2)} = 34.47, p < 0.001$ | $p < 0.001$ | $p < 0.001$ | $p = 0.01$ |
| PCL-R Factor 2 | 16.17 (1.69) | 11.43 (3.08) | 1.09 (1.44) | $H_{(2)} = 43.40, p < 0.001$ | $p < 0.001$ | $p < 0.001$ | $p < 0.001$ |
| PCL-R Facet 3 (lifestyle) | 7.59 (1.23) | 5.36 (1.69) | 1.00 (1.23) | $H_{(2)} = 41.07, p < 0.001$ | $p < 0.001$ | $p < 0.001$ | $p < 0.001$ |
| PCL-R Facet 4 (antisocial) | 8.47 (1.33) | 6.07 (2.13) | 0.50 (1.14) | $H_{(2)} = 41.84, p < 0.001$ | $p < 0.001$ | $p < 0.001$ | $p = 0.002$ |
| PCL-R Total | 28.21 (3.22) | 17.51 (4.55) | 2.64 (2.97) | $H_{(2)} = 45.55, p < 0.001$ | $p < 0.001$ | $p < 0.001$ | $p < 0.001$ |
| PD other than ASPD, N (%) |  |  |  |  |  |  |  |
| Cluster A | 5 (29%) | 0 (0) | . | $4.91, p = 0.05$ † | . | . | . |
| Cluster B | 8 (47%) | 2 (14%) | . | $3.77, p = 0.07$ † | . | . | . |
| Cluster C | 1 (6%) | 2 (14%) | . | $0.62, p = 0.58$ † | . | . | . |
| Lifetime SUD, N (%) | 3 (18%) | 4 (29%) | . | $0.52, p = 0.67$ † | | | |
| ADHD, N (%) | 2 (12%) | 1 (7%) | . | $0.19, p = 1.00$ † | | | |
| Positive urine drug test, N (%) |  |  |  |  |  |  |  |
| Placebo scan | 11 (65%) | 3 (23%) | 5 (23%) | $8.13, p = 0.02$ † | $p = 0.01$ | $p = 1.00$ | $p = 0.03^{\diamond}$ |
| Oxytocin scan | 12 (71%) | 5 (36%) | 6 (27%) | $\chi^2 = 7.78, p = 0.02$ | $p = 0.01$ | $p = 0.72$ | $p = 0.08$ |

### Group, treatment, and interaction effects on rCBF

The whole-brain analysis revealed a significant main effect of group in five clusters (Table 2, Figure 1). Post-hoc pairwise comparisons showed that both ASPD+P and ASPD-P had reduced rCBF relative to non-offenders in four of these clusters. These four clusters spanned the right hemisphere frontal and temporal areas (medial superior frontal gyrus, anterior cingulate cortex, orbitofrontal cortex, Rolandic operculum, pre-/post-central gyrus, and superior temporal gyrus). By contrast, in the fifth cluster, which encompassed some core areas of the posterior DMN (right hemisphere posterior cingulate, precuneus, hippocampus), both ASPD groups had increased rCBF relative to non-offenders, and ASPD+P had further increased rCBF compared to the ASPD-P. There were no significant main effects of treatment (oxytocin). However, a significant group by treatment interaction effect was found in one cluster spanning the left basal ganglia, specifically the globus pallidus, putamen and caudate (Table 2, Figure 2). Simple main effects tests revealed that this was driven by a significant decrease in rCBF after OT in ASPD-P only. These results were largely unchanged after adding substance use (presence of a positive drug screen) as a covariate (see supplementary materials).

**Table 2.**
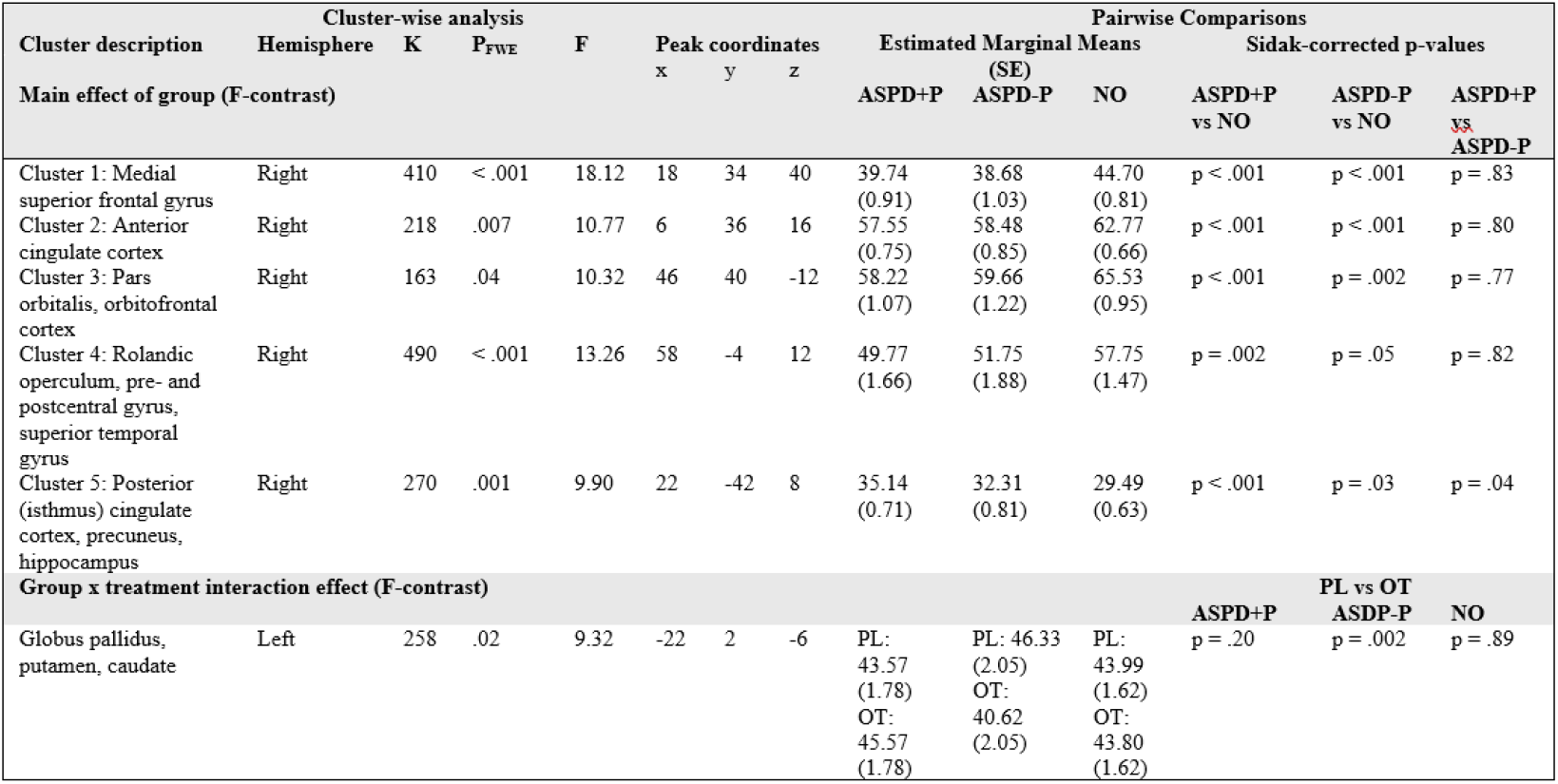
Clusters with significant group and interaction effects on rCBF. Note: main effects were measured with F-contrasts. PL = placebo scan, OT = oxytocin scan. Clusters labelled according to Automated Anatomical Labelling (AAL3) atlas built into SPM12 and confirmed by mapping MNI peak coordinates to Talairach space in BioImage Suite (https://bioimagesuiteweb.github.io/bisweb-manual/tools/mni2tal.html). SE = standard error.

**Figure 1.**
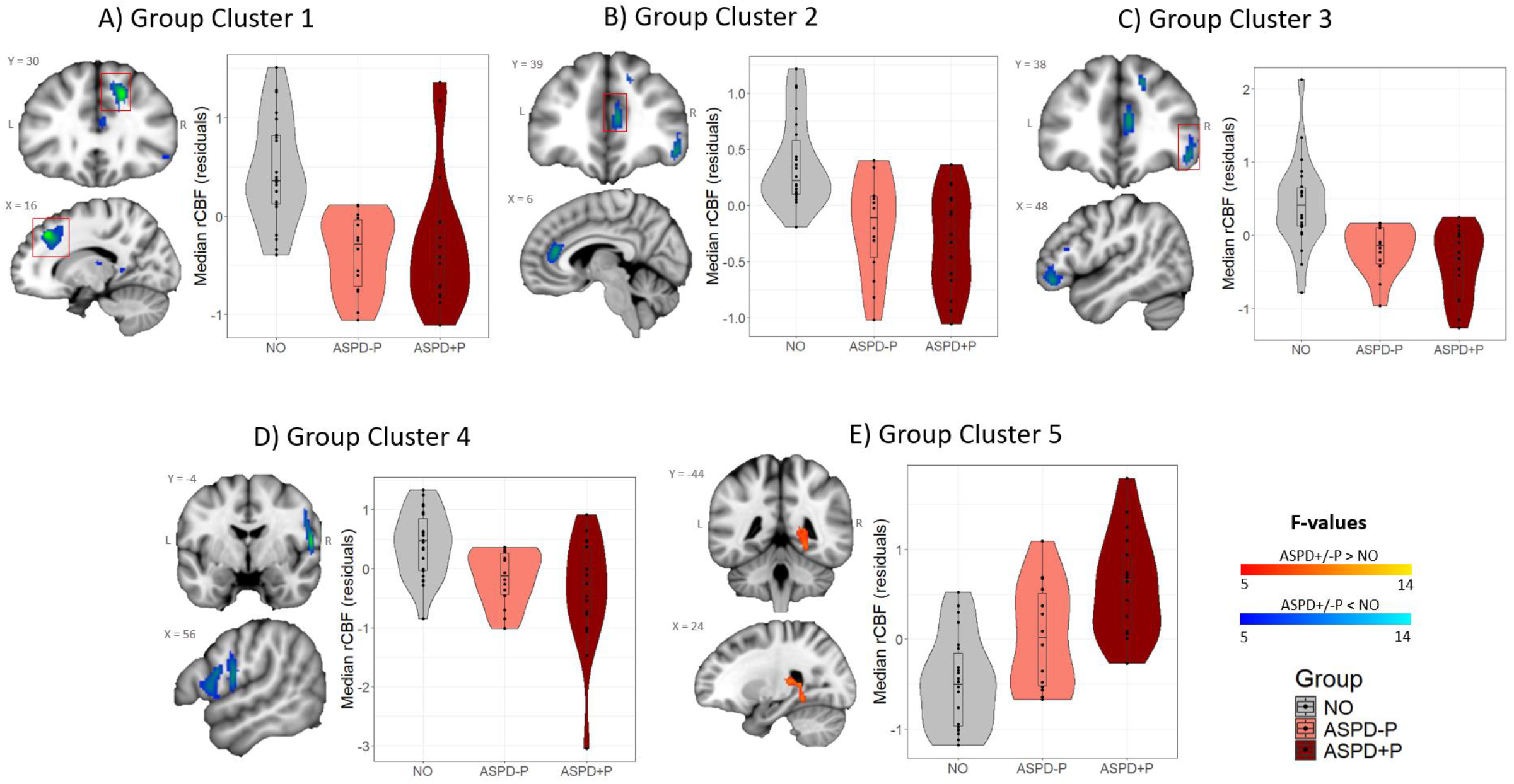
Clusters with significant group differences in rCBF. Note: The violin plots (with box plots inside) show the marginal mean and individual datapoints (z-standardized residuals) to depict spread of the median rCBF values within each significant cluster, and to clarify post-hoc what was driving the F-effects identified in the whole-brain analysis. For D), the main effect of group and the pairwise comparisons remained significant (p = 0.008) after removing the ASPD+P outlier (z-standardized residual > |3.0|), so this was kept in the analysis to increase power. Blue shaded clusters indicate reductions, red shaded cluster indicates increase. A red box visually highlights the cluster that the plot is referring to (in case other clusters are also visible in that slice).

**Figure 2.**
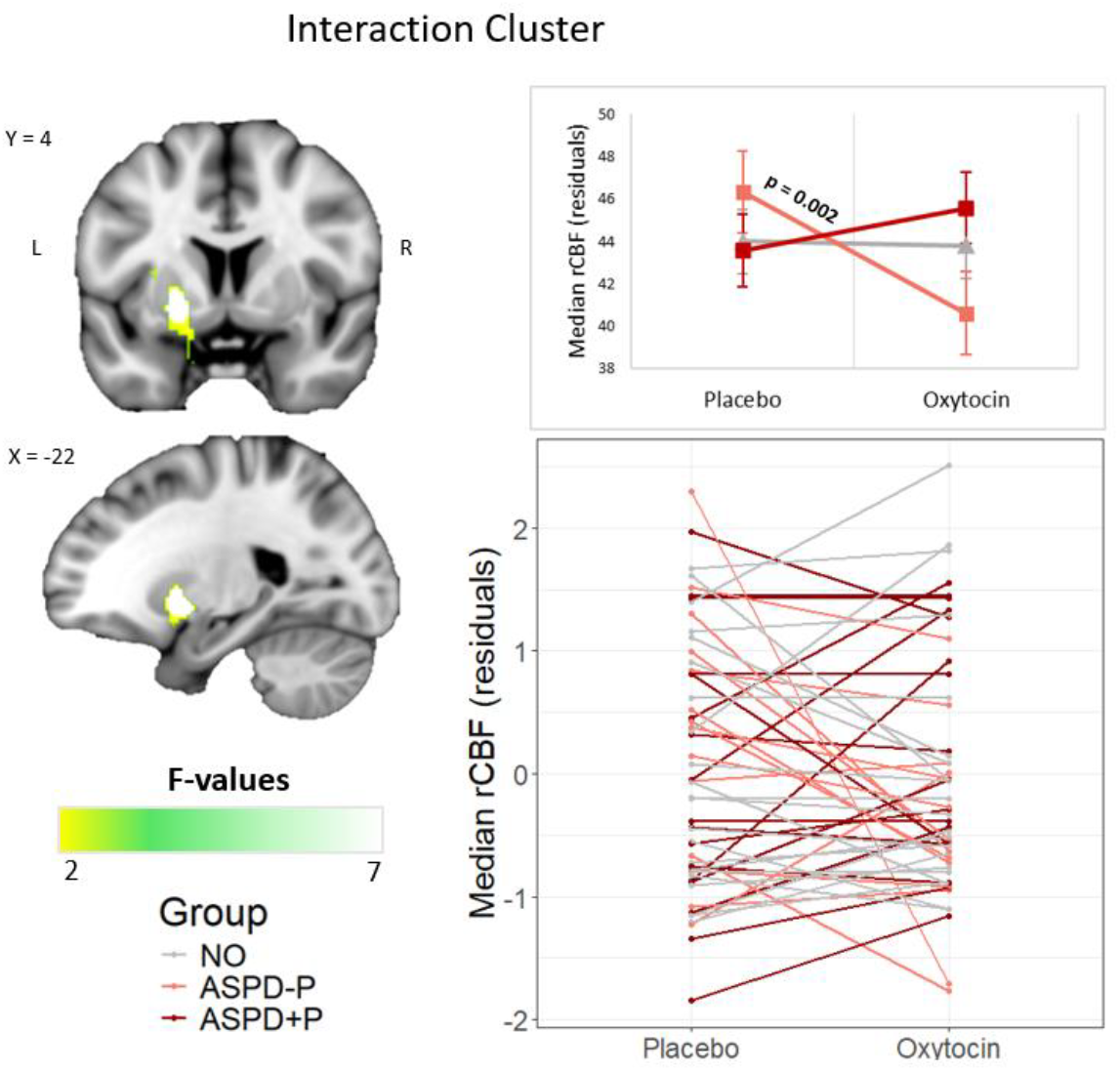
Cluster with a significant group by treatment interaction effect on rCBF. Note: The top line plot shows the marginal means (EMMs) of the median rCBF values for each group under each treatment condition, after accounting for the effect of global median CBF, age, and minutes since dose. The bottom spaghetti plot shows individual participants’ responsivity to OT. The interaction effect and simple main effect remain significant (p = 0.01) after excluding the single ASPD-P participant with the steepest slope.

The ROI analysis in the amygdala and anterior insula did not reveal significant group, treatment, or interaction effects (see supplementary table 2). The additional Bayesian linear mixed models revealed strong evidence in favour of the null hypothesis (see supplementary results). This indicated that there is a very low likelihood for group differences or treatment effects (or their interaction) on rCBF in the four ROIs.

The exploratory correlations with phenotypic characteristics did not yield significant results after correction for multiple comparisons (see supplementary table 3).

## Discussion

This study provides novel evidence that violent offenders with ASPD+P and ASPD-P exhibit shared and distinct resting-state neurobiological features. Both subtypes showed reduced rCBF in frontotemporal regions compared to non-offenders, while the ASPD+P group demonstrated higher rCBF in posterior DMN regions compared to the ASPD-P and non-offender groups. Intranasal oxytocin selectively reduced resting-state rCBF in the left basal ganglia of the ASPD-P group but had no significant effect in the ASPD+P or non-offender groups. These findings suggest that ASPD+P and ASPD-P have distinct neurobiological and neurochemical mechanisms, which should inform tailored therapeutic interventions.

In the whole-brain analysis, we observed reductions in rCBF in both ASPD+P and ASPD-P groups compared to non-offenders in frontotemporal regions. These are functionally implicated in the disordered cognitive processes common to both violent offender groups, such as learning from punishment cues, changing behaviour in the face of changing contingencies, and optimal decision-making processes [5]. Reduced resting-state rCBF [29,30] and BOLD activity [13–15,59–61] has been previously documented in these areas.. However, the key advantages in our approach were that ASL is more spatially exact and non-invasive than SPECT and PET scanning approaches, and our clinical population was more precisely phenotyped. The observed rCBF disruptions exist at rest, suggesting an intrinsic neurobiological dysfunction that may impair recruitment of these regions during cognitive tasks [23]. Reduced resting-state rCBF implies lower oxygen and nutrient delivery, potentially impacting the cognitive processes subserved by the affected regions. However, as ASL primarily measures perfusion, such behavioural implications remain speculative. Future studies could integrate cerebral metabolic rate of oxygen consumption (CMRO_2_) or oxygen extraction fraction (OEF) [62,63] measurements to determine whether these rCBF reductions reflect metabolic deficits in ASPD subtypes beyond the resting-state.

This study’s precise phenotypic characterization enabled us to determine the relative contribution of psychopathy to the resting-state brain function of individuals with ASPD. The ASPD+P group demonstrated significantly increased rCBF in a medial parietal cluster compared to the ASPD-P and non-offender groups. Previous studies of ASPD+P have consistently found grey [11] and white matter [64] structural, functional [13,36], and connectivity [65] abnormalities in this region, especially in the posterior cingulate and precuneus. Increased grey matter volume, surface area and BOLD demands may underpin the observed higher regional CBF. Yet there may also be important functional explanations. Typically, this region has heightened perfusion in the resting state, reflecting its role as a ‘rich club’ hub in the brain’s information processing network [20,66]. However, alterations in network topology in psychopathy [26] could make increased function in areas like the precuneus a compensatory mechanism which underpins information integration. Furthermore, this region contributes to functions including self-/other-referential processing (including mentalizing and perspective-taking), autobiographical episodic memory, and subjective reward representation [66–70]. Evidence suggests that automatic mentalizing and subject reward representational processes which draw on such areas are compromised in individuals with ASPD+P [13,71–76].

The medial parietal region is also a core part of the DMN, which displays brain activity when the individual is awake and alert but not actively engaged in an attention-demanding task [66,77]. DMN activity typically reduces during task engagement, but it has been shown that the posterior DMN does not appropriately deactivate during task engagement in psychopathic offenders [78]. This finding may be explained by neurodevelopmental processes, as it has recently been shown that functional networks such as the DMN are marked by distinct cellular fingerprints [79], which have genetic origins that may in turn be altered in individuals with psychopathy. While the relationship between aberrant brain function and gene expression requires further investigation in psychopathy, it has been established in other neurodevelopmental disorders such as autism [80].

We did not establish the expected resting-state rCBF abnormalities in the anterior insula or amygdala in the ASPD+P group. This supports recent findings which provide a more nuanced view of the role of functional amygdalar impairments in psychopathy’s aetiology, intrinsic baseline abnormality the role of amygdala impairments in psychopathy’s aetiology, further suggesting that amygdala dysfunction in psychopathy is context-dependent and task-evoked [81], rather than an intrinsic baseline abnormality [23]. Collaborative neuroimaging consortiums such ENIGMA-Antisocial Behaviour will explore the robustness of such negative findings by employing within-subject multimodal data with further improved statistical power (https://enigma.ini.usc.edu/ongoing/enigma-antisocial-behavior/).

Resting-state rCBF responsivity to oxytocin also differed between the ASPD groups. Thus, OT significantly reduced rCBF in the left basal ganglia, specifically the globus pallidus and dorsal striatum (putamen and caudate), when compared to placebo, in the ASPD-P but not the ASPD+P group. Importantly, this result is unlikely to reflect non-specific vascular effects, as OT (10-40 IU) does not disrupt cerebrovascular reactivity [42].

The basal ganglia, rich in oxytocin receptors [82], are crucial for reinforcement learning, reward processing, habit formation, and goal-directed action selection and control [83–85]. The dorsal striatum predominantly mediates choice impulsivity, evaluating action-contingent outcomes to better select future goal-directed actions [86]. Functional MRI studies have provided evidence for dorsal striatum abnormalities in antisocial groups in childhood [87]. Youths with disruptive behaviour disorders (precursors of ASPD in adulthood) show reduced responsiveness to positive prediction errors and increased responsiveness to negative prediction errors within the dorsal striatum during feedback [88] and reduced dorsal striatal response to early stimulus-reinforcement exposure [89]. Therefore, striatal dysfunction may underpin dysfunctional learning and decision-making in antisocial populations. Given the modulatory effect of OT on dorsal striatum function in this and other research [43,90], and evidence for a beneficial effect of OT on reinforcement learning [91–93], further studies investigating the potential therapeutic relevance of OT for reinforcement learning and decision-making in adult ASPD offenders are warranted.

We observed no significant OT effects on resting-state function in the ASPD+P group. One possibility is that ASPD+P individuals have higher peripheral endogenous OT levels [94,95], potentially limiting the impact of exogenous administration ([96] though see [97]). Alternatively, spatial pharmacodynamic studies show differential effects of OT on rCBF at different post-dose intervals [42,43], and it is possible that effects in ASPD+P may have been more immediate than in ASPD-P and thus not captured within our time window of 85 minutes post-dose. For example, OT effects in the amygdala, insula, and anterior cingulate might have been captured at an earlier time. These speculations emphasize the need for further studies sampling the effects of different post-dose intervals, different dose responses [41] and alternative administration routes on neural responsivity in ASPD+/-P.

Several limitations warrant consideration. First, OT effects were measured 85 minutes post-dose, which is later than most fMRI studies ([36,42,43] which have demonstrated a functional impact of OT in antisocial populations. Second, our study was limited to male participants, precluding conclusions about potential sex differences in OT responsivity [98]. Third, we did not demonstrate significant correlations with phenotypic characteristics of the ASPD+/-P groups, perhaps reflecting an underpowered approach [99]. Finally, while we observed significant OT effects in ASPD-P, individual variability in neural responses suggests the need for personalized treatment approaches. Larger trials incorporating genetic and endocrine markers of OT signalling may help to further enhance precision in ASPD treatment strategies.

In conclusion, this study using ASL to measure rCBF discovered novel evidence that ASPD+/-P have both shared and distinct resting-state functional neurobiological and neurochemical underpinnings. This adds support to the notion of stratifying ASPD into more biologically homogenous clinical subgroups for both mechanistic and therapeutic studies. Furthermore, it provided novel evidence that OT modulates rCBF in ASPD-P individuals, which lends support to the further exploration of OT as a potential therapeutic agent in personalised medicine approaches to the disorder.

## Data availability statement

All data are available from the authors upon reasonable request.

## Acknowledgements

Thank you to Dr Eleanor Hind for supporting with data collection. Thank you to the probation officers, especially David Bryan, for facilitating most offender recruitment.

## Funding

Funding for the research study was provided by Wellcome Clinical Research Training Fellowship grant for JT (grant no. 200099/S/15/S). Additional funding and financial support of the research team (JG, DMa, DMu, NG) was provided by the National Institute for Health and Care Research (NIHR) Maudsley Biomedical Research Centre (BRC) and an Economic and Social Research Council (ESRC) grant to YP (grant no. ES/K009400/1). The views expressed are those of the author(s) and not necessarily those of the Wellcome Trust, ESRC, NIHR or the Department of Health and Social Care.

## Competing interests

The authors declare no competing interests.

## Notes

### Competing Interest Statement

The authors have declared no competing interest.

### Clinical Trial

NCT05383300

### Author Declarations

The London City and East Research Ethics Committee (15/LO/1083) and the National Offender Management Services Research Committee (2016-382) gave ethical approval for this work.

