## Supplementary Materials for "Resting-state brain function and its modulation by intranasal oxytocin in antisocial personality disorder with and without psychopathy"

### Assessment procedure

Supplementary figure 1 shows the schedule and protocol of a typical scanning day appointment. The timing for the imaging protocol was largely adhered to, though in 2 ASPD+P participants, the task-based fMRI scans were omitted due to time pressure for both placebo and oxytocin scans, bringing the ASL scan closer in time to the spray administration. The starting time (+/- 10 minutes) was also largely adhered to, however, it shifted slightly for some participants due to scanner or individual availability changes, with 1 NO participant starting the oxytocin imaging protocol at 14:19 and another at 12:38 (and placebo at normal schedule), 1 ASPD-P participant starting the placebo imaging protocol at 09:53 and another at 12:37 (and oxytocin at normal schedule), 1 ASPD-P participant starting the oxytocin imaging protocol at 12:52 (and placebo at normal schedule), 1 ASPD+P participant starting the placebo imaging protocol at 15:38 and the oxytocin imaging protocol at 16:20, another starting the placebo imaging protocol at 16:38 and the oxytocin imaging protocol at 12:38, and a third starting the placebo imaging protocol at 09:52 and the oxytocin imaging protocol at 09:01 (the latter two being the participants that also skipped the task-based fMRI scans). Differences in timing since spray administration were accounted for across analyses, though within-subject differences in start time were not.

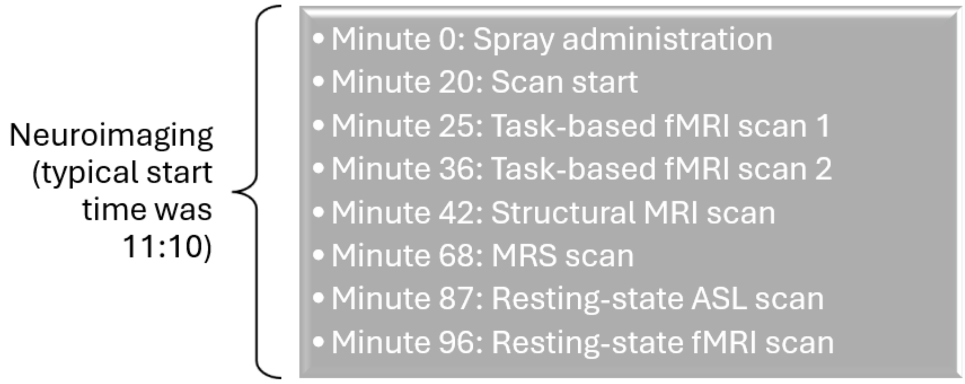


Supplementary figure 1. Overview of the schedule and planned imaging protocol for scanning days.

### Bayesian linear mixed models

As a post hoc assessment of non-significant results in the ROI analyses, we used R to conduct Bayesian linear mixed models, to compare these to the corresponding null models, and to calculate the Bayes Factor ($BF_{01}$) which provides insight into the strength of the evidence for the null hypothesis. Here is an example of the code for the right amygdala:

```
RHAmyg.fullmodel <- brm(Median ~ (Group*Substance) +
GlobalMedian_meancentred + Age_meancentred +
MinutesSinceFinalPuff_meancentred + (1|OXYASP_ID), data=RHAmyg.data,
family = gaussian(), prior = c(set_prior("normal(0,10)", class =
"b")), iter = 4000, warmup = 1000, chains = 4, cores = 4, control =
list(adapt_delta = 0.95), save_pars = save_pars(all = TRUE))
```

```
RHAmyg.nullmodel <- brm(Median ~ GlobalMedian_meancentred +
Age_meancentred + MinutesSinceFinalPuff_meancentred + (1 |
OXYASP_ID), data = RHAmyg.data, family = gaussian(), iter = 4000,
```

```
warmup = 1000, chains = 4, cores = 4, control = list(adapt_delta =
0.95), save_pars = save_pars(all = TRUE))

RHAmygBF <- bayes_factor(RHAmyg.fullmodel, RHAmyg.nullmodel)

RHAmygBF01 <- 1 / RHAmygBF$bf
```

### Supplementary results

#### Global median CBF

There was no significant main effect of group (F(2, 48) = 0.53, p = .59, $\eta_p^2$ = .02), treatment (F(1, 48) = 1.23, p = .27, $\eta_p^2$ = .03), or group by treatment interaction effect (F(2, 48) = 1.93, p = .16, $\eta_p^2$ = .07) on global median CBF (supplementary table 1).

| Global Median CBF | ASPD+P | ASPD-P | NO | Main effect of group | Main effect of treatment | Group x treatment |
|---|---|---|---|---|---|---|
| PL, mean (SD) | 47.59 (9.53) | 46.07 (11.06) | 44.77 (7.02) | F(2, 48) = 0.53, p = .59 | F(1, 48) = 1.23, p = .27 | F(2, 48) = 1.93, p = .16 |
| OT, mean (SD) | 46.35 (7.93) | 41.50 (7.74) | 46.36 (8.45) | | | |

Supplementary table 1. Global median CBF mean and standard deviation (SD).

#### ROI analysis

The boot-strapped linear mixed models revealed no significant group, treatment, or group by treatment interaction effect on rCBF in the amygdala or the anterior insula after FDR correction for multiple comparisons (supplementary table 2). Prior to correction, there was a significant treatment effect in the left insula (uncorrected p = 0.02). The covariate of no-interest global median CBF had a significant effect on all areas, and the covariate of no-interest age had a significant effect on right and left amygdala. Minutes since dose did not have a significant effect.

| Effect | Test statistic | FDR-corrected p | Effect size ($\mu_p^2$) |
|---|---|---|---|
| **Right amygdala** | | | |
| Group | $F_{(2, 47.61)} = 1.72$ | 0.39 | 0.07 |
| Treatment | $F_{(1, 49.06)} < 0.01$ | 0.98 | < 0.001 |
| Group x treatment | $F_{(2, 49.70)} = 0.90$ | 0.74 | 0.03 |
| Global median CBF | $F_{(1, 96.79)} = 538.31$ | <0.001 | 0.85 |
| Age | $F_{(1, 48.26)} = 6.27$ | 0.04 | 0.10 |
| Minutes since dose | $F_{(1, 90.47)} = 0.83$ | 0.48 | 0.01 |
| **Left amygdala** | | | |
| Group | $F_{(2, 48.61)} = 1.55$ | 0.39 | 0.06 |
| Treatment | $F_{(1, 49.77)} = 0.31$ | 0.77 | 0.01 |
| Group x treatment | $F_{(2, 50.41)} = 1.72$ | 0.74 | 0.06 |
| Global median CBF | $F_{(1, 93.81)} = 478.55$ | <0.001 | 0.84 |
| Age | $F_{(1, 49.28)} = 8.75$ | 0.02 | 0.15 |
| Minutes since dose | $F_{(1, 95.43)} = 0.05$ | 0.83 | < 0.001 |
| **Right insula** | | | |
| Group | $F_{(2, 47.81)} = 1.33$ | 0.39 | 0.05 |
| Treatment | $F_{(1, 48.11)} = 0.68$ | 0.77 | 0.01 |
| Group x treatment | $F_{(2, 48.62)} < 0.01$ | 0.99 | < 0.001 |
| Global median CBF | $F_{(1, 73.75)} = 569.89$ | <0.001 | 0.89 |
| Age | $F_{(1, 48.39)} = 0.07$ | 0.80 | 0.002 |
| Minutes since dose | $F_{(1, 88.97)} = 1.96$ | 0.35 | 0.02 |
| **Left insula** | | | |
| Group | $F_{(2, 47.88)} = 1.06$ | 0.39 | 0.04 |
| Treatment | $F_{(1, 48.77)} = 5.34$ | 0.08* | 0.10 |
| Group x treatment | $F_{(2, 49.38)} = 0.61$ | 0.74 | 0.02 |
| Global median CBF | $F_{(1, 88.33)} = 569.70$ | <0.001 | 0.87 |
| Age | $F_{(1, 48.55)} = 0.35$ | 0.80 | 0.007 |
| Minutes since dose | $F_{(1, 97.00)} = 3.94$ | 0.21 | 0.04 |

Supplementary table 2. ROI analysis of rCBF in amygdala and anterior insula. * Uncorrected p = 0.02.

To examine the non-significant results further, we conducted Bayesian linear mixed models. The full model, which included the fixed effects of group, treatment, and their interaction, alongside covariates (global median CBF, age, and minutes since dose) as well as the random effect of subject was compared to the null model, which only included the covariates and the random effect of subject. For the right amygdala, Bayes Factor ($BF_{01}$) was 780772148, for the left amygdala, $BF_{01}$ = 249713017, for the right insula, $BF_{01}$ = 261256027, and for the left insula, $BF_{01}$ = 29610173. Across all four ROIs, this suggests that there is compelling evidence in favour of the null hypothesis, indicating that group, treatment, or their interaction do not significantly influence median rCBF after adjusting for the covariates. This validates the results from the frequentist linear mixed model.

### Correlation with phenotype

Exploratory bootstrapped partial correlations across all ASPD participants did not reveal any significant relationship between phenotypic characteristics and median rCBF in areas with significant group or interaction effects. However, as indicated in supplementary table 3, there were two correlations that were significant prior to correction for multiple comparisons, namely a positive correlation between the presence of a reconviction within 3 years of participation and rCBF in cluster 1 (i.e., the right medial superior frontal gyrus where ASPD participants showed reduced rCBF compared to non-offenders), as well as a positive correlation between proactive aggression and rCBF in cluster 4 (the right Rolandic operculum, pre-/post-central gyrus, superior temporal gyrus, where ASPD also showed reduced rCBF compared to non-offenders). Although these correlations may not be in an expected direction, the results nonetheless encourage further exploration of the relationship between rCBF and phenotypic characteristics of ASPD and psychopathy within larger samples.

| | Group effect | | | | | Interaction |
|---|---|---|---|---|---|---|
| | Cluster 1 | Cluster 2 | Cluster 3 | Cluster 4 | Cluster 5 | effect |
| # violent convictions | -.022 | .028 | .291 | -.058 | .027 | .015 |
| Reconviction within 3 years | **.484*** | .076 | .280 | -.290 | -.094 | .013 |
| Reactive aggression | .104 | -.018 | -.026 | .212 | -.256 | .110 |
| Proactive aggression | .152 | -.180 | .094 | **.425*** | -.075 | .090 |
| PCL-R factor 1 | -.059 | -.252 | .015 | -.010 | .160 | .273 |
| PCL-R factor 2 | -.008 | -.133 | -.060 | -.096 | -.200 | -.004 |

Supplementary table 3. Correlation between rCBF findings and phenotypic characteristics across all ASPD participants (for aggression measures, N = 26 as not all 31 ASPD participants completed the questionnaire). * Uncorrected p < 0.05.

### Effect of substance use

Sensitivity analyses were conducted to assess the effect of recent substance use as indicated by a positive urine drug test from the day of each MRI scan. Specifically, this binary variable was included as an additional covariate (alongside age, global rCBF, and time since administration) in the post-hoc tests that were used to interpret the main effect F-contrasts.

The results remained largely the same as without including this variable as covariate. Only the ASPD-P vs NO comparison in cluster 4 lost significance. All other results remained the same or became more significant.
